# Admission biomarkers may not have an association with cardiovascular involvement in Latin American patients with Multisystem Inflammatory Syndrome in Children (MIS-C)

**DOI:** 10.1101/2023.06.13.23291351

**Authors:** Jose M Galindo-Hayashi, Karen González-Moctezuma, Oscar Tamez-Rivera, Ana Victoria Villarreal-Treviño

**Author notes:** **Corresponding author and first author:** Jose M Galindo-Hayashi. All authors have seen and approved the manuscript.

## Abstract

**Objectives:** The aim of the study is to determine if there is an association between altered biomarkers and cardiovascular involvement in Latin American patients with MIS-C.

**Design:** The researchers of this study conducted a retrospective cohort study.

**Setting:** Secondary care maternal unit hospital in Monterrey, NL, Mexico.

**Participants:** Any register of a Latin-American pediatric patient with MIS-C within the database from March 2019 to February 2022.

**Primary and secondary outcome measures:** Cut, mean, odds ratio (OR), relative ratio (RR), 95% confidence interval (CI) and *p* values of inflammatory markers associated with cardiovascular affection in MIS-C. As a secondary outcome we measure being male as an independent risk factor.

**Results:** None of the biomarkers and gender variables taken were significant (Table 1).

**Conclusions:** The researchers’ analysis suggests there is no evidence of cause-effect association between admission biomarkers and the presence of cardiovascular affection in MIS-C. Remarkably, neutrophilia and ESR had a high odds ratio and a nearly significant p-value, which makes them ideal for further analysis with a bigger sample. Additionally, gender was included as a risk factor and was analyzed independently, nonetheless, it was not associated with a higher risk of presenting cardiovascular affection.

**Article summary:** 

**Strengths and limitations of this study:**

- First study measuring the association of a series of available and economical biomarkers with a higher incidence of cardiovascular involvement in MIS-C in a Latin-American population.
- Set ground for further research regarding the current treatment of MIS-C
- Sample size non-representative.
- Since this is an observational study we can’t fully control confounding and bias (e.j. reporting bias)

## Introduction

Multisystem Inflammatory Syndrome in Children (MIS-C) is an uncommon but serious complication of SARS-CoV-2 infection that has been responsible for approximately 2% of the pediatric deaths attributed to COVID-19. Although MIS-C and Kawasaki Disease (KD) exhibit similar presenting symptoms, their inflammatory pathways and clinical outcomes differ. The exact pathophysiology of MIS-C remains unknown; however, it has been theorized that this post-infectious inflammatory condition occurs in part due to a hyperimmune response of the host (1, 2).

Previous and current diagnostic criteria for MIS-C require the presence of fever and laboratory evidence of inflammation, as well as multisystem organ involvement (1, 16). Studies suggest that patients with MIS-C exhibit higher levels of C-reactive protein (CRP), troponin, erythrocyte sedimentation rate (ESR), serum ferritin, procalcitonin, and D-dimer compared to patients with other inflammatory and infectious diseases. (2, 3, 8, 12, 13, 16, 17, 18). Cardiovascular involvement is a core feature of MIS-C that has been reported in >80% of the patients; and it may present as shock, myocarditis, pericarditis, low ventricular ejection fraction (<55%), congestive heart failure, mitral valve regurgitation, and coronary artery dilation/aneurysms (1, 6, 7, 10, 13). Likewise, cardiovascular involvement in patients with MIS-C has been associated with an increased risk of admission to the intensive care unit (ICU) and a higher mortality risk (1, 2, 10).

Recent studies have focused on the characterization of the host’s immune response in the context of MIS-C in order to establish possible biomarkers that predict organ involvement and clinical outcomes. A recent study in Latin American children with MIS-C found that the elevation of multiple cytokines (CXCL8, CXCL9, CXCL10, IL-6, IL-10, IL-18) is associated with pericardial effusion and shock (20).

In spite of these interesting findings, the availability of most of these biomarkers may be limited in certain clinical scenarios. Furthermore, the availability of known and widely accepted cardiovascular biomarkers such as troponins, brain natriuretic peptide (BNP), and N-terminal proBNP (NT-proBNP) may be scarce in developing and resource-limited countries. Considering that hispanic patients are disproportionately affected by MIS-C (21), the study of readily available biomarkers that may predict cardiovascular involvement is required. We aimed to evaluate possible surrogate and readily available biomarkers associated with cardiovascular involvement in children with MIS-C in a limited-resource clinical scenario.

## Methods

### Design

Retrospective, unicentric, observational and analytic cohort study.

### Setting and population analyzed

The participants were selected from the database of a secondary maternal unit in Monterrey, NL, Mexico. The eligibility criteria were Latin-American pediatric patients with MIS-C treated from March 2019 to February 2022.

We used the WHO MIS-C criteria to choose the eligible patients to analyze.

The criteria considered to diagnose cardiovascular involvement were ETT findings of: pericarditis, EKG abnormalities, valvular regurgitation, pericardial effusion, coronary aneurysm, myocarditis, ventricular failure. The ETT was performed prior to the administration of IVIG and/or steroids.

### Statistics - sampling, measuring, testing, analyzing

The researchers dichotomize MIS-C patients in those with and without cardiovascular involvement. SPSS and excel statistical software’s were used to define statistical significance. The researchers performed descriptive statistics to analyze demographic characteristics in both groups.

The researchers analyzed inflammatory markers using an odds ratio test to find association to cardiovascular involvement. Furthermore, a chi-square test was used to test statistical significance. The following markers were analyzed: C-reactive protein (CRP), erythrocyte sedimentation rate (ESR), Ferritin, D-dimer, creatine kinase (CK), leukocytes, neutrophils, lymphocytes, lactate dehydrogenase (LDH), platelets and fibrinogen. The cut and p values are represented in the following table (table 1).

**Table 1.**

| Risk factor | Association between risk factor (inflammatory markers and gender) and outcome (with or without cardiovascular affection) |
| --- | --- |
| **Leukocytosis** (Cut value >15,500 /mm<sup>3</sup>) | **Mean:** 13,247 /mm<sup>3</sup><br>**OR:** 1.72<br>**RR:** 1.35<br>**p:** 0.31 |
| **Neutrophilia** (Cut value >8,000 /mm<sup>3</sup>) | **Mean:** 10,429 /mm<sup>3</sup><br>**OR:** 2.52<br>**RR:** 1.73<br>**p:** 0.08 |
| **CRP** (Cut value >0.5mg/dL) | **Mean:** 14 mg/dL<br>**OR:** 2.25<br>**RR:** 1.71<br>**p:** 0.48 |
| **ESR** (Cut value >15 mm/hr) | **Mean:** 25 mm/hr<br>**OR:** 3.27<br>**RR:** 2.25<br>**p:** 0.08 |
| **Hyperferritinemia** (Cut value >1,000 ng/dL) | **Mean:** 2,918 ng/dL<br>**OR:** 1.22<br>**RR:** 1.12<br>**p:** 0.13 |
| **D-dimer** (Cut value 2,000 mg/dL) | **Mean:** 7,628 mg/dL<br>**OR:** 3.25<br>**RR:** 2.24<br>**p:** 0.14 |
| **Fibrinogen** (Cut value >515 mg/dL) | **Mean:** 669 mg/dL<br>**OR:** 1.38<br>**RR:** 1.2<br>**p:** 0.56 |
| **Lymphopenia** (Cut value <2,000 /mm3) | **Mean:** 1,919 /mm3<br>**OR:** 1.02<br>**RR:** 1.01<br>**p:** 0.96 |
| **Thrombocytopenia** (Cut value <150,000 /m3) | **Mean:** 193,098 /m3<br>**OR:** 1.12<br>**RR:** 1.07<br>**p:** 0.82 |
| **LDH** (Cut value >220 U/L) | **Mean:** 449 U/L<br>**OR:** 1.91<br>**RR:** 1.51<br>**p:** 0.45 |
| **CK** (Cut value >200 U/L) | **Mean:** 964 U/L<br>**OR:** 1.01<br>**RR:** 1.01<br>**p:** 0.03 |
| **Hypoalbuminemia** (Cut value <3.5 g/dL) | **Mean:** 4 g/dL<br>**OR:** 1.14<br>**RR:** 1.08<br>**p:** 0.8 |
| **Male** | **RR:** 0.9<br>**OR:** 0.84<br>**p:** 0.76 |

The laboratory exams were taken at the moment of admission, prior to any intervention (corticosteroids or IVIG). *OR=Odds ratio, RR: Relative ratio, CI: 95% confidence interval, p= p value, CRP: C-Reactive protein, ESR: Erythrocyte sedimentation rate, CK: Creatine Kinase, LDH: Lactate dehydrogenase.*

### Primary outcome measures

Cut, mean, odds ratio (OR), relative ratio (RR), 95% confidence interval (CI) and p values of inflammatory markers associated with cardiovascular affection in MIS-C.

### Secondary outcome measures

OR, RR and p value of being male as a risk factor associated with cardiovascular affection in MIS-C.

### Patient and Public Involvement

Patients or the public were not involved in the design, or conduct, or reporting, or dissemination plans of our research.

## Results

70 patients fulfilled the eligibility criteria, six were excluded for not having performed an ETT, four were excluded for having an incomplete file. 60 patients were analyzed.

The demographic characteristics were the following. 35 were men (58%), 25 were women (42%), 25 patients had cardiovascular affection verified by ETT (42%). We found a higher prevalence of cardiovascular affection in men (56%). The mean age in patients with MIS-C and cardiovascular affection was 7 years and 1 month.

The cut value, mean, odds ratio (OR), relative ratio (RR), 95% confidence interval (CI) and p values of inflammatory markers associated with cardiovascular affection in MIS-C are represented in table 1.

## Discussion

The association of cardiovascular affection in MIS-C and morbidity has been reported worldwide in large-scale studies (1, 6, 7). The literature reports that patients with MIS-C and cardiac involvement are at a higher risk of ICU admission (2, 8).

Inflammatory and cardiac markers have been linked to a higher association of cardiovascular affection, such as aspartate aminotransferase, thrombocytopenia, BNP and/or troponin T. However, the link is still not well established (2, 8, 10, 12, 14). Furthermore, in Mexico some of these markers are not easily available, and if they are, they usually are at a high cost (BNP and troponin). A panel of specialists with expertise in MIS-C, concluded that some patients with mild symptoms may require only close monitoring without immunomodulatory treatment (4, 15). So, it is plausible to infer, a valid treatment option would be exclusive use of IVIG and/or steroids when there is evidence of inflammatory markers associated with cardiovascular affection and/or clinically unstable patients.

The researchers’ purpose for this study was to identify which, easily available, biomarkers are associated with cardiovascular involvement in MIS-C among a Latin-American pediatric population. This is the first study measuring the association of a series of available and economical inflammatory markers with a higher incidence of cardiovascular involvement in MIS-C in a Latin-American population.

The researchers’ analysis reported it is plausible that there is a cause-effect association between a high value of leukocytes (Cut value >15,500 /mm3) and the presence of cardiovascular affection in MIS-C, with a p value of 0.001 (95% C.I.: 10,836.1 - 15,657.9) which suggests it is unlikely to be associated by chance. Additionally, the odds ratio was 10.66 which suggests the patients are 10 times more likely to present cardiovascular affection in MIS-C when leukocytes at admission are >5,500 /mm3. However, an odds ratio with a sample this size is mildly significant. A significant p value was obtained with CK (p = 0.03); however, the sample size was small (n=47), which resulted in a wide confidence interval (0 - 2,269), which makes it of poor statistical value. The rest of inflammatory markers were non-significant. The researchers’ highlight neutrophilia and ESR which had a high odds ratio and a nearly significant p-value, which makes them ideal for further analysis with a bigger sample. Additionally, gender was included as a risk factor and was analyzed independently, nonetheless, it was not associated with a higher risk of presenting cardiovascular affection (p=0.76).

Nevertheless, since this is an observational study, we can’t fully control confounding and bias (e.j. reporting bias) and therefore we suggest ultimately to design a randomized blinded trial to prove this association. With the final purpose of prognosticating which patients would benefit the most from IVIG and/or steroid therapy in centers with limited resources.

## Data Availability

The data presented in this study are available upon reasonable request from the corresponding author. The data is not publicly available due to privacy concerns.

## Author Contributions

1. Jose M Galindo-Hayashi. **Contribution**: Conception and design, acquisition of data, analysis and interpretation, revising the article. (First and corresponding author)

2. Karen González-Moctezuma. **Contribution**: Acquisition of data, analysis and interpretation, revising the article.

3. Oscar Tamez-Rivera. **Contribution:** Analysis and interpretation, revising the article.

4. Ana Victoria Villarreal-Treviño. **Contribution**: Revising the article, design, analysis, and interpretation.

## Acknowledgments

We thank José Guillermo Rodriguez, MD, and Regina Ayala for assistance in data collection and management.

## Limitations

Bias and confounding are issues since it is an observational study.

Confounders (Age, severity, comorbidity, echocardiography skills of the cardiologist, accuracy of the diagnosis) Bias (reporting bias-overdiagnosis of MIS-C)

## Transparency declaration

The lead author affirms that this manuscript is an honest, accurate, and transparent account of the study being reported; that no important aspects of the study have been omitted; and that any discrepancies from the study as planned (and, if relevant, registered) have been explained.

## Ethics approval

The study was conducted according to the guidelines of the Declaration of Helsinki and approved by the Research Ethics Committee of the “Hospital Regional Materno Infantil de Alta Especialidad” with registration number DEISC-PR-19 01 21 065.

## References

1. National Center for Immunization and Respiratory Diseases. Health Department-Reported Cases of Multisystem Inflammatory Syndrome in Children (MIS-C) in the United States. Cdc [Internet]. 2020;1–7. Available from: https://www.cdc.gov/mis-c/cases/index.html

2. Pignatelli R, Antona CV, Rivera IR, Zenteno PA, Acosta YT, Huertas-Quiñones M, et al. Pediatric multisystem SARS COV2 with versus without cardiac involvement: a multicenter study from Latin America. European Journal of Pediatrics. 2021;180(9):2879–88.

3. Abrams JY, Oster ME, Godfred-Cato SE, Bryant B, Datta SD, Campbell AP, et al. Factors linked to severe outcomes in multisystem inflammatory syndrome in children (MIS-C) in the USA: a retrospective surveillance study. The Lancet Child and Adolescent Health [Internet]. 2021;5(5):323–31. Available from: http://dx.doi.org/10.1016/S2352-4642(21)00050-X

4. McCrindle BW, Rowley AH, Newburger JW, Burns JC, Bolger AF, Gewitz M, et al. Diagnosis, treatment, and long-term management of Kawasaki disease: A scientific statement for health professionals from the American Heart Association. Vol. 135, Circulation. 2017. 927–999 p.

5. Son, M. B. F., Murray, N., Friedman, K., Young, C. C., Newhams, M. M., Feldstein, L. R., Loftis, L. L., Tarquinio, K. M., Singh, A. R., et al. Overcoming COVID-19 Investigators (2021). Multisystem Inflammatory Syndrome in Children - Initial Therapy and Outcomes. The New England journal of medicine, 385(1), 23–34. https://doi.org/10.1056/NEJMoa2102605

6. Swann, O. V., Holden, K. A., Turtle, L., Semple, M. G., et al. (2020). Clinical characteristics of children and young people admitted to hospital with covid-19 in United Kingdom: prospective multicentre observational cohort study. BMJ, m3249. https://doi.org/10.1136/bmj.m3249

7. Sokunbi, O., Akinbolagbe, Y., Akintan, P., et al. (2022). Clinical presentation and short-term outcomes of multisystemic inflammatory syndrome in children in Lagos, Nigeria during the COVID-19 pandemic: A case series. EClinicalMedicine, 49, 101475. https://doi.org/10.1016/j.eclinm.2022.101475

8. Zhao, Y., Patel, J., Huang, Y., Yin, L., & Tang, L. (2021). Cardiac markers of multisystem inflammatory syndrome in children (MIS-C) in COVID-19 patients: A meta-analysis. The American Journal of Emergency Medicine, 49, 62–70. https://doi.org/10.1016/j.ajem.2021.05.044

9. Budnik O, I., Hirsch B, T., Fernández C, C., Yánez P, L., & Zamorano R, J. (2011). Enfermedad de Kawasaki: una serie clínica. Revista Chilena De Infectología, 28(5), 416–422. https://doi.org/10.4067/s0716-10182011000600005

10. Alvarez Z., P., Larios G., G., Toro R., L., et al. (2020). Recomendación para la sospecha, manejo y seguimiento del compromiso cardiovascular en pacientes con Síndrome Inflamatorio Multisistémico en Pediatría asociado a COVID-19 (PIMS-TC). Declaración de Posición de la Sociedad Chilena de Pediatría (SOCHIPE) y Sociedad Chilena de Cardiología y Cirugía Cardiovascular (SOCHICAR). Revista Chilena De Pediatría, 91(6), 982. https://doi.org/10.32641/rchped.v91i6.3215

11. Lima-Setta, F., Magalhães-Barbosa, M. C. D., Rodrigues-Santos, Prata-Barbosa, A. et al. (2021). Multisystem inflammatory syndrome in children (MIS-C) during SARS-CoV-2 pandemic in Brazil: a multicenter, prospective cohort study. Jornal De Pediatria, 97(3), 354–361. https://doi.org/10.1016/j.jped.2020.10.008

12. Capone, C. A., Subramony, A., Sweberg, T., et al. (2020). Characteristics, Cardiac Involvement, and Outcomes of Multisystem Inflammatory Syndrome of Childhood Associated with severe acute respiratory syndrome coronavirus 2 Infection. TheJournal of Pediatrics, 224, 141–145. https://doi.org/10.1016/j.jpeds.2020.06.044

13. Kohli, U., & Lodha, R. (2020). Cardiac Involvement in Children With COVID-19. Indian Pediatrics, 57(10), 936–940. https://doi.org/10.1007/s13312-020-1998-0

14. Ganguly, M., Nandi, A., Banerjee, P., Gupta, P., Sarkar, S. D., Basu, S., & Pal, P. (2021). A comparative study of IL-6, CRP and NT-proBNP levels in post-COVID multisystem inflammatory syndrome in children (MISC) and Kawasaki disease patients. International Journal of Rheumatic Diseases, 25(1), 27–31. https://doi.org/10.1111/1756-185x.14236

15. Licciardi, F., Baldini, L., Dellepiane, M., et al. (2021). MIS-C Treatment: Is IVIG Always Necessary? Frontiers in Pediatrics, 9. https://doi.org/10.3389/fped.2021.753123

16. Melgar, M., Lee, E. H., Miller, A. D., Lim, et al. (2022). Council of State and Territorial epidemiologists/CDC surveillance case definition for multisystem inflammatory syndrome in children associated with SARS-COV-2 infection — United States. MMWR. Recommendations and Reports, 71(4), 1–14. https://doi.org/10.15585/mmwr.rr7104a1

17. Kostik, M. M., Bregel, L. V., Avrusin Chasnyk, V. G. (2022). Heart Involvement in Multisystem Inflammatory Syndrome, Associated With COVID-19 in Children: The Retrospective Multicenter Cohort Data. Frontiers in Pediatrics, 10. https://doi.org/10.3389/fped.2022.829420

18. Chan, J. F. W., Yuan, S., Kok, K. H., Yuen, K. Y. (2020). A familial cluster of pneumonia associated with the 2019 novel coronavirus indicating person-to-person transmission: a study of a family cluster. The Lancet, 395(10223), 514–523. https://doi.org/10.1016/s0140-6736(20)30154-9

19. Alsaied, T., Tremoulet, A. H., Burns, J. C., Saidi, A., Dionne, A., Lang, S. M., Newburger, J. W., de Ferranti, S., & Friedman, K. G. (2021). Review of Cardiac Involvement in Multisystem Inflammatory Syndrome in Children. Circulation, 143(1), 78–88. https://doi.org/10.1161/circulationaha.120.049836

20. Lambert, K., Moo, K. G., Arnett, A., Goel, G., Hu, A., Flynn, K. J., Speake, C., Wiedeman, A. E., Gersuk, V. H., Linsley, P. S., Greenbaum, C. J., Long, S. A., Partridge, R., Buckner, J. H., & Khor, B. (2022). Deep immune phenotyping reveals similarities between aging, Down syndrome, and autoimmunity. Science translational medicine, 14(627), eabi4888. https://doi.org/10.1126/scitranslmed.abi4888

21. Jiang, L., Tang, K., Levin, M., Irfan, O., Morris, S. K., Wilson, K., Klein, J. D., & Bhutta, Z. A. (2020). COVID-19 and multisystem inflammatory syndrome in children and adolescents. The Lancet. Infectious diseases, 20(11), e276–e288. https://doi.org/10.1016/S1473-3099(20)30651-4

